# Topo-Net: Retinal Image Analysis with Topological Deep Learning

**DOI:** 10.1101/2024.02.03.24302291

**Authors:** Faisal Ahmed, Baris Coskunuzer

## Abstract

The analysis of fundus images for the early screening of eye diseases is of great clinical importance. Traditional methods for such analysis are time-consuming and expensive as they require a trained clinician. Therefore, the need for a comprehensive and automated clinical decision support system to diagnose and grade retinal diseases has long been recognized. In the past decade, with the substantial developments in computer vision and deep learning, machine learning methods have become highly effective in this field to address this need. However, most of these algorithms face challenges like computational feasibility, reliability, and interpretability.

In this paper, our contributions are two-fold. First, we introduce a very powerful feature extraction method for fundus images by employing the latest topological data analysis methods. Through our experiments, we observe that our topological feature vectors are highly effective in distinguishing normal and abnormal classes for the most common retinal diseases, i.e., Diabetic Retinopathy (DR), Glaucoma, and Age-related Macular Degeneration (AMD). Furthermore, these topological features are interpretable, computationally feasible, and can be seamlessly integrated into any forthcoming ML model in the domain. Secondly, we move forward in this direction, constructing a topological deep learning model by integrating our topological features with several deep learning models. Empirical analysis shows a notable enhancement in performance aided by the use of topological features. Remarkably, our model surpasses all existing models, demonstrating superior performance across several benchmark datasets pertaining to two of these three retinal diseases.

## I. Introduction

The World Health Organization reports that as of 2019, more than 400 million people worldwide suffer from Glaucoma, diabetic retinopathy (DR), age-related macular degeneration (AMD), or other serious eye diseases [1]. As most patients with eye diseases are not aware of the aggravation of these conditions, early screening and treatment of eye diseases are quite important. Currently, detecting these conditions is a time-consuming and manual process that requires a trained clinician to examine and evaluate digital color fundus images of the retina, which can result in delayed treatment. Therefore, the need for clinical decision-support methods has long been recognized.

With this motivation, machine learning (ML) methods have been widely employed in retinal image analysis in the past decade [2], [3]. These efforts have made substantial progress in the field by using image classification and pattern recognition [4]. Especially after the success of convolutional neural networks (CNN) in image classification, ML tools proved to be quite effective in retinal image analysis [5]. However, these methods are not either computationally efficient to work in large datasets or interpretable to provide insights to ophthalmologists for disease diagnosis.

In this paper, we bring a novel approach towards retinal disease diagnosis, introducing topological data analysis (TDA) methods to fundus imaging. Over recent years, TDA tools have garnered success in medical image analysis across various domains, adeptly capturing concealed shape patterns within images and generating powerful feature vectors from these patterns (Sec. II). Despite its application in numerous biomedical domains [6], this potent approach has not been previously utilized within the retinal image analysis domain. In our work, we identify that retinal images exhibit distinct topological patterns with varying color values, such as numerous small loops and large loops of differing sizes Figure 1. Employing the key method in TDA, persistent homology, we transform these patterns into topological feature vectors suitable for utilization with any ML method. In this research, we developed two ML models, *Topo-ML* and *Topo-Net*, using these feature vectors. The first model, Topo-ML, shows high computational feasibility, managing very large datasets in mere hours, while the second, Topo-Net, represents a great integration of current deep learning models with these topological features. Utilizing these models, we investigated the diagnosis of the three most common retinal diseases: Diabetic Retinopathy (DR), Glaucoma, and Age-related Macular Degeneration (AMD). While our computationally feasible Topo-ML stands shoulder to shoulder with state-of-the-art deep learning models on benchmark datasets, our Topo-Net model outperforms them in DR and AMD diagnosis by a substantial margin (Sec. V-C).

**Fig. 1:**
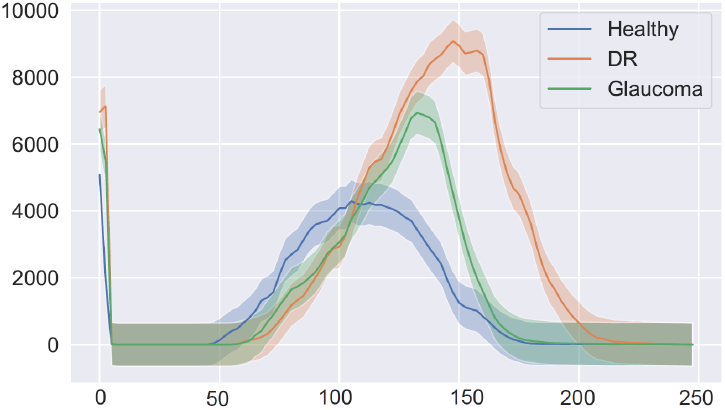
Our topological feature vectors effectively differentiate between various diseases. Above, we present median curves and 40% confidence bands for the Betti-1 vectors on the HRF dataset.

### Our contributions

- We bring a new perspective to retinal image analysis by introducing TDA methods to the field.
- We extract topological features of fundus images for most common retinal diseases (DR, Glaucoma, and AMD) and observe easily detectable and *interpretable* topological pattern differences (See Figure 1 and Section VI).
- Utilizing our topological feature vectors with conventional ML techniques, we first create a computationally efficient *Topo-ML model*, yielding highly competitive results across all diseases. Next, by integrating our topological features with the latest deep learning models, our *Topo-Net model* surpasses all state-of-the-art models in the DR and AMD benchmark datasets.
- Our topological features provide a valuable contribution to the field of retinal image analysis, offering seamless integration with any future ML model to boost their performance.

## II. Related Work

### Machine Learning in Retinal Image Analysis

In the past decade, ML tools have been widely employed in medical image analysis [7], [8]. Particularly, in the scope of retinal image analysis, ML methodologies have demonstrated notable effectiveness [5]. There are two mainstream applications of ML tools for retinal image analysis. The first is the diagnosis and grading of diseases which can be considered a classification problem for image data [9]. The second mainstream application is lesion detection/segmentation [2]. In this paper, we focus only on the diagnosis and grading of retinal diseases by using TDA methods.

Following the triumph of convolutional neural networks (CNN) in image classification tasks, deep learning methods proved to be quite effective in retinal image analysis [3], [10]. There is extensive literature for deep learning methods in ophthalmology, where excellent reviews can be found in the following recent surveys [2], [4], [11], [12].

### Topological Data Analysis in Image Processing

TDA stands as a novel methodology for investigating complex data by analysing both local and global patterns across various scales, thus addressing challenges tied to data dimensionality, variance in data collection methods, and changing scales [13], [14].

Over the past decade, TDA has been successfully applied across various domains such as image analysis, neurology, cardiology, hepatology, gene-level and single-cell transcriptomics, drug discovery, evolution, and protein structural analysis [15]. By tapping into the inherent topological features present in images, TDA brings a new perspective to image analysis. Its ability to uncover hidden image patterns forges new pathways for undertakings like image segmentation, object recognition, image registration, and image reconstruction [16].

A frequently utilized tool from TDA in image analysis is Persistent Homology (PH), which has manifested remarkable outcomes in pattern recognition for image and shape analysis over the span of two decades [17]. In the realm of medical image analysis, PH has displayed its effectiveness in the analysis of images related to hepatic lesions [18], histopathology [19], [20], fibrin images [21], tumor classification [22], Chest X-ray image screening [23], neuronal morphology [24], brain artery trees [25], fMRI data [26], [27], and genomics data [28]. A thorough review of TDA methodologies within biomedicine is encapsulated in the exemplary survey [6].

## III. Methodology

In this paper, we use *persistent homology* (PH) as a powerful feature extraction tool for retinal images. PH is one of the key approaches in topological data analysis (TDA), allowing us to systematically assess the evolution of various hidden patterns in the data as we vary a scale parameter [13], [29]. The extracted patterns, or homological features, along with information on how long such features persist throughout the considered filtration of a scale parameter, convey a critical insight into salient data characteristics and data organization. In this section, we give a basic introduction to PH in image data settings, which is called *cubical persistence*. For a thorough background and PH process for other data types (e.g., point clouds, networks), see [30], [31].

### A. PH for Image Data: Cubical Persistence

PH can be summarized as a 3-step process. The first step is the *filtration* step, where one induces a sequence of simplicial complexes from the data. This is the key step, where one can integrate the domain information into the process. The second step is the *persistence diagrams*, where the machinery records the evolution of topological features (birth/death times) in this sequence of the simplicial complexes. The final step is the *vectorization* where one can convert these records to a function or vector to be used in suitable ML models. For more details on how to apply PH in image analysis, check out the references given in Section II.

#### 1) Constructing Filtrations

As PH is basically the machinery to keep track of the evolution of topological features in a sequence of simplicial complexes, the most important step is the construction of this sequence. In the case of image analysis, the most common method is to create a nested sequence of binary images (aka cubical complexes). For a given image *𝒳* (say *r × s* resolution), to create such sequence, one can use grayscale (or other color channels) values *γ*_*ij*_ of each pixel Δ_*ij*_ ⊂ *𝒳* . In particular, for a sequence of grayscale values (*t*_1_ < *t*_2_ < *· · ·* < *t*_*N*_ ), one obtains nested sequence of binary images *𝒳* _1_ ⊂ *𝒳* _2_ ⊂ *· · ·* ⊂ *𝒳* _*N*_ such that *𝒳* _*n*_ = *{*Δ_*ij*_ ⊂ *𝒳* | γ_*ij*_ ≤ *t*_*n*_} (See Figure 2 and 3). In other words, we start with empty *r × s* image and start activating (coloring black) pixels when their grayscale value reaches the given threshold. This is called *sublevel filtration* for *𝒳* with respect to a given function (grayscale in this case). One can also go in decreasing order to activate the pixels, which is called *superlevel filtration*.

**Fig. 2:**
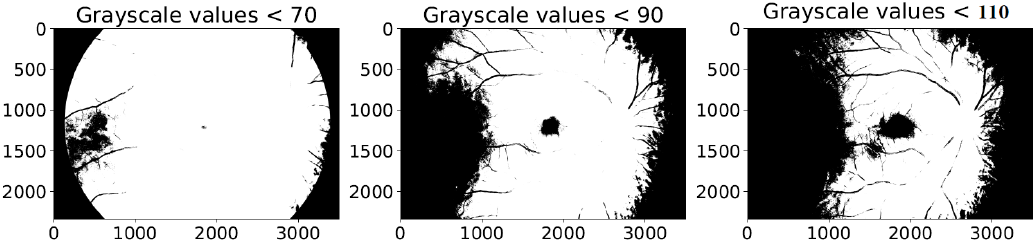
Sublevel filtration. Binary images *𝒳* _70_, *𝒳* _90_, *𝒳* _110_ obtained from a fundus image for threshold values 70, 90, 110.

**Fig. 3:**
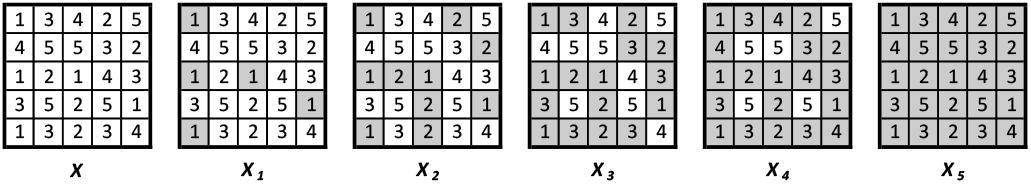
Toy sublevel filtration. The leftmost figure represents an image of 5 *×* 5 size with the given pixel values. Then, the sublevel filtration is the sequence of binary images *𝒳*_1_ ⊂ *𝒳*_2_ ⊂ *· · ·* ⊂ *𝒳*_5_.

The choice of thresholds is also very crucial in this construction as it indicates how fine we want our PH machinery to detect the topological patterns. One can choose *N* = 255 which makes the filtration too fine, and most outputs would be trivial in the fingerprinting process. If we choose *N* very small, then one can miss many topological features. While in most cases, the thresholds are chosen evenly distributed, depending on the image dataset, using quantiles of the grayscale values is also common, e.g., when grayscale values in the dataset are concentrated around specific numbers. In our experiments, we use *N* = 100 as we obtain very good results with this choice, and increasing N did not improve the performance significantly.

Note that sublevel and superlevel filtration produces completely different information for most data types. However, in the image data setting (cubical complexes), in the complementary dimensions, sublevel and superlevel filtration produce very similar information thanks to a celebrated result in algebraic topology, Alexander Duality [32]. In other words, the choice of sublevel and superlevel filtration is not important in image data as long as one uses all possible dimensions (in this setting, *k* = 0, 1).

#### 2) Persistence Diagrams

The second step in PH process is to obtain persistence diagrams (PD) for the filtration *𝒳* _1_ ⊂ *𝒳* _2_ ⊂ *· · ·* ⊂ *𝒳* _*N*_, i.e., the sequence of cubical complexes (binary images). PDs are formal summaries of the evolution of topological features in the filtration sequence. PDs are collection of 2-tuples, *{*(*b*_*σ*_, *d*_*σ*_)}, marking the birth and death times of the topological features appearing in the filtration. In other words, if a topological feature *σ* appears for the first time at 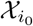, we mark the birth time *b*_*σ*_ = *i*_0_. Then, if the topological feature *σ* disappears at 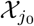, we mark the death time *d*_*σ*_ = *j*_0_. i.e., PD_*k*_ (*𝒳*) = {(*b*_*σ*_, *d*_*σ*_) | *σ* ∈ 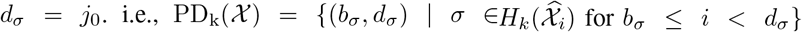 for *b*_*σ*_ ≤ *i* < d_*σ*_ }. Here, 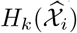 represent *k*^*th*^ homology group of 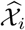, representing k-dimensional topological features in cubical complex 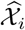 [32]. By construction, for 2*D* image analysis, only meaningful dimensions to use are *k* = 0, 1, i.e., PD_0_(*𝒳* ) and PD_1_(*𝒳* ).

In layman’s terms, 0-dimensional features are connected components and 1-dimensional features are the holes (loops). In our case, if a loop τ first appears at the binary image *𝒳*_3_ and it gets filled in the binary image *𝒳*_7_, we add 2tuple (3, 7) in the persistence diagram PD_1_(*𝒳* ). Similarly, if a new connected component appears in the binary image *𝒳*_5_ and it merges to the other components in the binary image *𝒳*_8_, we add (5, 8) to PD_0_(*𝒳* ). In Figure 3, we have PD_0_(*𝒳* ) = *{*(1, ∞), (1, 2), (1, 3), (1, 3), (1, 4), (2, 3)} and PD_1_(*𝒳* ) = *{*(3, 5), (3, 5), (4, 5)} This step is pretty standard and there are various software libraries for this task. For image data with cubical complexes, see [33]. For other types of data and filtrations, see [34].

#### 3. Vectorization

PDs being a collection of 2-tuples are not very practical to be used with ML tools. Instead, a common way is to convert PD information into a vector or a function, which is called *vectorization* [35]. A common function for this purpose is the *Betti function*, which basically keeps track of the number of “alive” topological features at the given threshold. In particular, the Betti function is a step function with β_0_(*t*_*n*_) the count of connected components in the binary image *𝒳*_*n*_, and β_1_(*t*_*n*_) the number of holes (loops) in *𝒳*_*n*_. In ML applications, Betti functions are usually taken as a vector 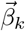 of size N with entries β_*k*_(*t*_*n*_) for 1 ≤ *n* ≤ *N*.

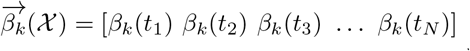

For example, for the image *𝒳* in Figure 3, we have 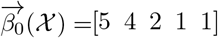 and 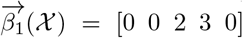, e.g., β_0_(1) = 5 is the count of components in *𝒳*_1_ and β_1_(4) = 3 is the count of holes (loops) in *𝒳*_4_. There are other vectorization methods like Persistence Images, Persistence Landscapes, or Silhouettes [30], but to keep our model interpretable, we use Betti functions in this study.

### B. Color Channels for Retinal Images

Retinal image quality assessment is essential for controlling the quality of retinal imaging and guaranteeing the reliability of diagnoses by ophthalmologists or automated analysis systems. The three main families of conventional color spaces are primary spaces, luminance-chrominance spaces, and perceptual spaces. Appropriate color spaces can help simplify some color computations that occur during the generation of images. In 1978, Joblov et al. [36] described the significance of different color spaces in computer graphics and the feature extraction process. In our persistent homology approach, the way we construct the filtration out of the given fundus image is the key step (Section III), and the different color channels induce completely different filtrations and produce different topological patterns.

### C. Extracting Topological Features

In this part, we first describe our topological fingerprinting machine learning model, Topo-ML. Then, we elaborate on the explainability and interpretability of our model.

In the flowchart (Figure 4), we summarized our Topo-ML model. For a given fundus image *𝒳* (say at *r × s* resolution), we first get its RGB and Grayscale images (Figure 4-Step 2). In other words, we produce 4 color functions **g**(*i, j*) (grayscale=average(RGB)), **R**(*i, j*) (red), **G**(*i, j*) (green), and **B**(*i, j*) (blue) where f(*i, j*) assigns every pixel Δ_*ij*_ ⊂ *𝒳* to its assigned color value for 1 ≤ *i* ≤ *r* and 1 ≤ *j* ≤ *s*. Note that all colors have a range [0, 255] . Then, we extract all topological features for color channel values by constructing a sublevel filtration with respect to the corresponding color function. While grayscale values vary from 0 (black) to 255 (white), we chose the number of thresholds as *N* = 100 in our filtration step, as further increasing the threshold steps did not increase the performance of our model. In other words, we renormalized [0, 255] grayscale interval to [0, 100]. After defining the filtration *𝒳*_1_ ⊂ *𝒳*_2_ ⊂ *· · ·* ⊂ *𝒳*_100_, we obtain the persistent diagrams PD_*k*_(*𝒳* ) of each fundus image *𝒳* for dimensions *k* = 0, 1 (Section III). In layman’s terms, the filtration produces a sequence of binary (black-white) images where the dark points represent the pixels value less than the given threshold (Figure 2). Then, PD_0_(*𝒳* ) records connected components in these binary images, and PD_1_(*𝒳* ) records holes in the binary images.

**Fig. 4:**
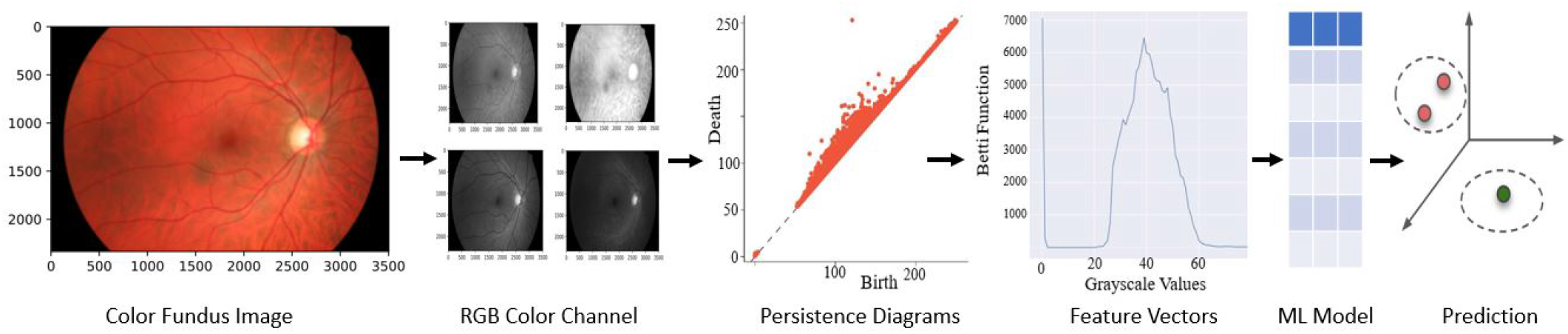
Flowchart of Topo-ML model. We start with fundus images, extract RGB and Gray color spaces, and then create persistence diagrams from color values. Topological feature vectors (Betti functions) are derived from these diagrams and fed into machine learning models like RF, XGBoost, and kNN, resulting in highly accurate classifications.

**Fig. 5:**
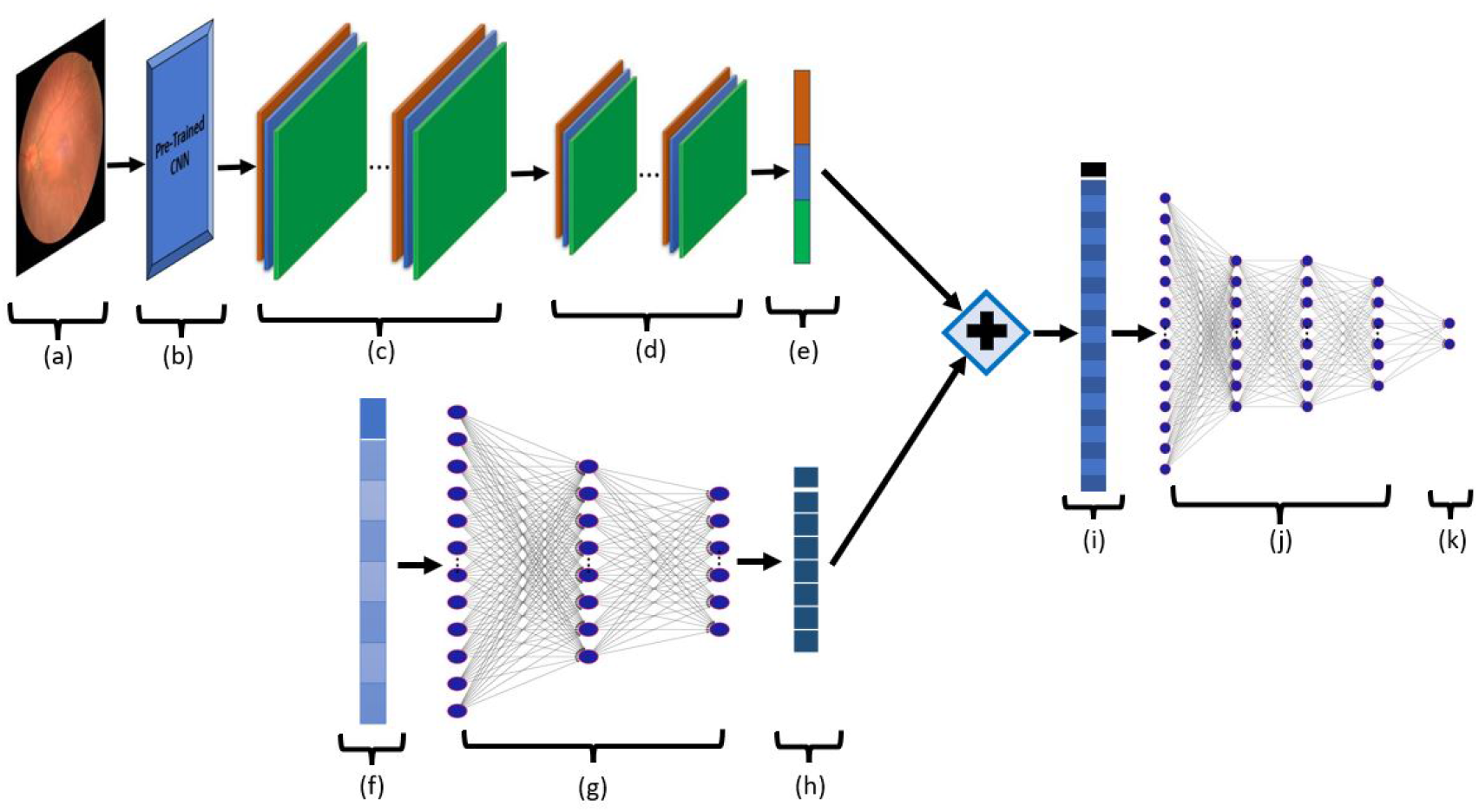
Architechture of Topo-Net model. Our model takes two inputs (a) fundus image and (f) topological feature vector of the fundus image. In the top row, we run our CNN layers: (b) Pre-trained model weights (c) convolution layer (d) Max pooling layer, and (e) Flatten. In the bottom row, we run (g) feed-forward network on topological features to effectively adapt them into our model. We then concatenate (+) the produced vectors (e) and (h) and obtain (i). We run another feed-forward network to get our predictions.

After getting persistent diagrams, we convert them into feature vectors as explained in Section III. In this vectorization step, one can use several choices like Betti functions, Silhouettes, or Persistence Images. Since most of the topological features have short life spans, Betti and Silhouette (with *p* ≤ 1) functions were the natural choice as they give the count of topological features at a given threshold. Our experiments verified this intuition and we obtained almost all the best results with Betti functions. Hence, to keep our model simple, we only use Betti functions as vectorizations (Fig. 4-Step 4).

## IV. ML Models

We created two models to utilize these topological features extracted from fundus images. In the first one, to keep the model feasible, and show the strength of our features, we use a simple ML classifier, and get our *Topo-ML Model*. In the second one, we integrate these topological features with deep learning models to boost their performance. We call these topology enhance deep learning models *Topo-Net Model*.

### A. Topo-ML Model

After obtaining our topological feature vectors, the final step is to apply an ML classifier to these vectors. To keep our model computationally feasible, we applied tree-based methods like Random Forest and XGBoost to our extracted topological features. Note that our topological feature vectors can easily be integrated with various deep learning models, too. We give the details of our ML steps in the experiments section. Note that topological features are invariant under rotation, flipping, and translation. This makes our model highly robust against noisy data. Furthermore, our model does not need any data augmentation and data pre-processing which makes our model computationally very feasible. Our experiments show that our model is highly successful in small datasets as well as large ones.

### B. Topo-Net Model

To combine our topological features, we used several backbones for our deep learning model.

#### Pre-trained CNN Models for Retinal Images

The proposed Convolutional Neural Network (CNN) architecture leverages the power of transfer learning, building upon the foundation of the renowned pre-trained CNN models like ResNet50, DenseNet201, MobileNetV2, EfficientNetB3, Xception, VGG19, InceptionV3, EfficientNetB0, and EfficientNetB2. Initially, the pre-trained model is loaded with weights and set to input the shape of images of dimensions (256, 256, 3) while excluding its top layers. To preserve the valuable knowledge encoded in the pre-trained weights, they are effectively frozen, ensuring that they remain unchanged during training. Following this, a convolutional layer with 64 filters, each employing a (3, 3) kernel and the rectified linear unit (ReLU) activation function, is introduced. Subsequently, a MaxPooling layer reduces spatial dimensions, enhancing the network’s ability to capture salient features. The flattened layer transitions the data into a one-dimensional form, paving the way for a dense layer with 64 units, further enriched by the ReLU activation function. To mitigate overfitting, a dropout layer with a 0.4 rate is added. Finally, the architecture culminates in an output layer, which is adaptable for binary or multi-class classification tasks. For binary classification, a single sigmoid-activated unit is employed, while for multi-class problems, a softmax activation function is used with multiple units. This CNN architecture embodies the fusion of established pre-trained knowledge and task-specific adaptability, forming a potent tool for retinal image classification.

We illustrate our architecture for Topo-Net Model in Figure 5. We give the details in five steps as follows:

##### 1) Step 1: CNN Feature Extraction

We employ a pre-trained model with weights initialized from ImageNet to capture image features. The input shape for this CNN is (256, 256, 3). To avoid updating the pre-trained weights, we set `include_top` to False. We follow this with a 2D convolutional layer consisting of 64 filters with a kernel size of (3, 3) and ReLU activation. Subsequently, a max-pooling layer with a (2, 2) pooling window is applied. The feature maps are then flattened. Finally, a dense layer with 64 units and a ReLU activation function is added to further refine the CNN features.

##### 2) Step 2: TDA Feature Extraction

The input layer for TDA features is flattened. An activation layer with ReLU activation is applied. Two dense layers follow, with 256 and 128 units, both utilizing ReLU activation functions.

##### 3) Step 3: Concatenation of Features

The extracted CNN and TDA features are concatenated to create a fused feature representation.

##### 4) Step 4: Further Feature Processing

Two additional dense layers are introduced with 256 and 128 units, both using ReLU activation functions. A dropout layer with a rate of 0.2 is inserted to prevent overfitting.

##### 5) Step 5: Output Layer

The final output layer is chosen based on the nature of the classification task.For binary classification, a sigmoid activation function is applied. For multiclass classification, a softmax activation function is used.

This concatenated architecture seamlessly combines the strengths of both CNN and TDA features, enabling the model to capture complex patterns in the data for improved classification performance.

## V. Experiments

### A. Datasets

To see the performance of our Topo-ML model for Glaucoma, DR, and AMD screening, we did several experiments on well-known benchmark datasets. We give the basic details of these datasets in Table I. Further details (resolution, camera, etc.) for all the datasets can be found in [2].

**TABLE I:**
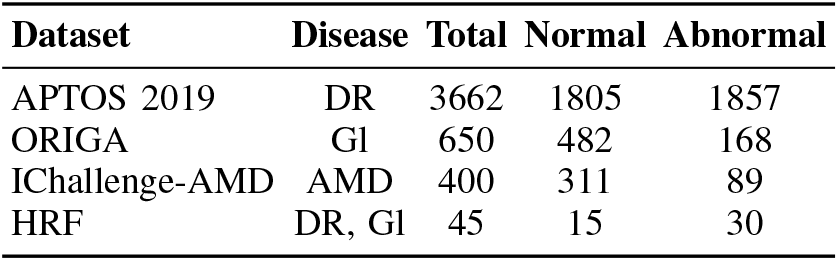
Benchmark datasets for fundus images.

**IChallenge-AMD dataset** is designed for the Automatic Detection challenge on Age-related Macular degeneration (ADAM Challenge) which was held as a satellite event of the ISBI 2020 conference [37], [38]. There are two different resolutions of images, i.e., 2124 *×* 2056 pixels (824 images) and 1444 *×* 1444 (376 images). While the dataset has 1200 images, only 400 of them are available with labels. Like most other references, we used these 400 images in our experiments Table II.

**TABLE II:** Accuracy results for AMD diagnosis.

| IChallenge-AMD Dataset |  |  |  |  |  |  |  |
| --- | --- | --- | --- | --- | --- | --- | --- |
| Method | Nor:Abn | Train:Test | Class | Prec | Recall | Acc | AUC |
| Mem-Bank [48] | 311:89 | 5 fold CV | 2 | 74.6 | 66.5 | 82.0 | 66.5 |
| Invariant [49] | 311:89 | 5 fold CV | 2 | 83.2 | 74.5 | 86.6 | 71.6 |
| Decouple [50] | 311:89 | 5 fold CV | 2 | 79.7 | 69.2 | 83.8 | 69.2 |
| Contrastive [51] | 311:89 | 5 fold CV | 2 | 73.5 | 68.1 | 82.5 | 68.1 |
| Rotation S [47] | 311:89 | 5 fold CV | 2 | 84.5 | 77.2 | 87.6 | 77.2 |
| CycleGAN [52] | 933:267 | 5 fold CV | 2 | 77.1 | 80.6 | 87.3 | 86.2 |
| DCNN [53] | 311:89 | 10 fold CV | 2 | 84.9 | 76.0 | 91.7 | 88.0 |
| **Topo-ML** | 311:89 | 80:20 | 2 | 88.9 | 57.1 | 91.3 | 90.6 |
| **Topo-Net** | 311:89 | 85:15 | 2 | **92.4** | **91.3** | **93.8** | **93.6** |

**ORIGA dataset** contains 650 high resolution (3072 *×* 2048) retinal images for Glaucoma annotated by trained professionals from Singapore Eye Research Institute [39].

**APTOS 2019 dataset** was used for a Kaggle competition on DR diagnosis [40]. The images have varying resolutions, ranging from 474 *×* 358 to 3388 *×* 2588. APTOS stands for Asia Pacific Tele-Ophthalmology Society, and the dataset was provided by Aravind Eye Hospital in India. In this dataset, fundus images are graded manually on a scale of 0 to 4 (0: no DR; 1: mild; 2: moderate; 3: severe; and 4: proliferative DR) to indicate different severity levels. The number of images in these classes are respectively 1805, 370, 999, 193, and 295. In the binary setting, class 0 is defined as the normal group, and the remaining classes (1-4) are defines as DR group which gives a split 1805:1857. The total number of training and test samples in the dataset were 3662 and 1928 respectively. However, the labels for the test samples were not released after the competition, so like other references, we used the available 3662 fundus images with labels.

### B. Experimental Setup

#### Training Test Split

Unfortunately, none of the datasets have a predefined *train:test* split, and therefore, many models used their own split. In our experiments, we used an 80:20 split in all datasets for Topo-ML model. For Topo-Net model, we used 80:20 split for IChallenge-AMD and ORIGA dataset, while for the APTOS dataset, we used 85:15 for binary and multi-class classifications which aligns more closely with the standard settings of previous research. Because of the discrepancy between the experimental setups of different methods, we give the train:test splits of all models in our accuracy tables to facilitate a fair comparison (Tables II to V).

#### Data Augmentation

Note that as our datasets are quite small and imbalanced compared to other image classification tasks for deep learning models, hence all CNN and other deep learning methods need to use serious data augmentation (sometimes 50-100 times) to train their model and avoid overfitting [41]. Our Topo-ML model are using topological feature vectors, and our feature extraction method is invariant under rotation, flipping and other common data augmentation techniques. Hence, we do not use any type of data augmentation or preprocessing to increase the size of training data for Topo-ML model. This makes our model computationally very efficient, and highly robust against small alterations and the noise in the image. Similarly, for Topo-Net model, we used the original datasets and did not use any kind of data augmentation as we used pre-trained models as backbone.

#### Topo-ML Model Hyperparameters

To increase the performance of our model in terms of accuracy and computational efficiency, we performed parametric tuning and feature selection methods. We extracted 800 features (Gray and RGB color spaces) from the datasets by using Betti 0 and Betti 1. To improve the performance and avoid collinearity, we used dimension reduction by choosing the most important features. For feature selection, we used *SelectFromModel* from scikitlearn. We first assign importance to each feature and sorted them in descending order according to threshold parameter. The features are considered unimportant and removed if the corresponding importance of the feature values is below the provided threshold parameter. Random Forest and XGBoost models are trained on all of the datasets. We used default parameters as parametric tuning for XGBoost. After feature selection and fine-tuning, XGBoost gave the best results for APTOS (159 features), ORIGA (84 features), and IC-AMD (58 features) datasets . As our ablation study (Table VI) indicates, feature selection improved both performance and computational time.

#### Topo-Net Model Hyperparameters

We trained the TopoNet model for 50 epochs using a batch size of 32 for all datasets. We used the Adam optimizer and kept the remaining parameters as default.

#### Computational Complexity & Implementation

While for high dimensional data PH calculation is computationally expensive [34], for image data, it is highly efficient. For 2D images, PH has time complexity of *𝒪* (|*𝒫*|^*r*^) where *r* ∼ 2.37 and |*𝒫*| is the total number of pixels [42]. In other words, PH computation increases almost quadratic with the resolution. The remaining processes (vectorization, RF) are negligible compared to PH step. We used Giotto-TDA [43] to obtain persistence diagrams, and Betti functions. We used Jupyter notebook as an IDE for writing the code in Python 3. Our code is available at the following link ^1^.

#### Runtime

We conducted all our experiments utilizing a personal laptop equipped with an Intel(R) Core(TM) i7-8565U processor running at 1.80GHz and boasting 16 GB of RAM. In both of our models, the most time-intensive phase is the extraction of topological features. In contrast, the subsequent tasks of machine learning classification and executing deep learning models are relatively insignificant. This is mainly because we did not employ any data augmentation and utilized pre-trained CNN models. For the most extensive dataset in our study, APTOS (consisting of 3662 high-resolution images), the entire process, including topological feature extraction, model training, and obtaining accuracy results, consumed a total of 43.7 hours. The runtime for smaller datasets with lower resolutions, used for topological feature extraction, is considerably shorter. It’s also worth considering that when using a server, as opposed to a personal laptop, the runtime for such datasets would be considerably shorter.

### C. Results

Here, we present the performance of our Topo-ML and Topo-Net models along with SOTA models on benchmark datasets. We give the accuracy results of our Topo-ML model for Glaucoma, AMD, and DR diseases. In Tables II to V, we give *Normal:Abnormal* split, *Train:Test* split, and # *Classes* to describe the experimental setting used for each baseline.

In these tables, we report all available performance metrics (AUC, Accuracy, Precision, Recall) provided with the baselines. For details of these performance metrics, see [44]. Our models are trained on the training data, and we report their performance on the test (unseen) data. Scikit-Learn [45] library is used for performance metric calculations. For multiclass classification (APTOS 2019), we used a weighted average of AUC values with a One-vs-Rest configuration. This method computes the AUC values of each class against the rest [46]. In all tables, the best result for each column is given in bold, and the second-best result is underlined. For missing data in the table from reference papers, we used “-”.

#### AMD Detection Results

Our results for diagnosing AMD on the IChallenge-AMD (IC-AMD) dataset are presented in Table II. The accuracy values listed for methods in rows 1-4 have been sourced from [47, Table 1]. While these papers did not directly employ the IC-AMD dataset, [47] adapted these methods to the IC-AMD dataset using the same experimental setup and subsequently reported these results. Although baseline results range from an AUC of 0.665 to 0.880, both of our models consistently outperform all existing models. The outstanding performance of our Topo-ML model, which solely relies on our topological vectors and XGBoost, underscores the effectiveness of our features in diagnosing AMD.

#### DR Detection Results

We give our results for DR diagnosis on APTOS 2019 dataset in binary setting in Table III and in multiclass setting in Table V. We gave the details for these settings in Section V-A. In binary setting, our basic model Topo-ML stands shoulder to shoulder with SOTA deep learning models. On the other hand, our Topo-Net model outperforms all existing models in all performance metrics. We note that because of the mixed resolution of the images, this is a very challenging dataset from ML perspective. In spite of this fact, both our models proved to be very robust and can tackle mixed-resolution problems.

**TABLE III:** Accuracy results for binary DR diagnosis.

| APTOS 2019 Dataset Binary (DR) |  |  |  |  |  |  |  |
| --- | --- | --- | --- | --- | --- | --- | --- |
| Method | Nor:Abn | Train:Test | Class | Prec | Recall | Acc | AUC |
| D-Net121 [54] | 1805:1857 | 85:15 | 2 | 86.0 | 87.0 | 94.4 | - |
| ConvNet [55] | 1805:1857 | 80:20 | 2 | - | - | 96.1 | - |
| DRISTI [56] | 1805:1857 | 85:15 | 2 | - | - | 97.1 | - |
| C-DNN [57] | 1805:1857 | 85:15 | 2 | 98.0 | 98.0 | 97.8 | - |
| LBCNN [58] | 1805:1857 | 80:20 | 2 | - | - | 96.6 | 98.7 |
| SCL [59] | 1805:1857 | 85:15 | 2 | 98.4 | 98.4 | 98.4 | **98.9** |
| **Topo-ML** | 1805:1857 | 80:20 | 2 | 95.5 | 94.0 | 94.5 | 97.9 |
| **Topo-Net** | 1805:1857 | 85:15 | 2 | **98.7** | **98.7** | **98.7** | **98.9** |

**TABLE IV:** Accuracy results for Glaucoma diagnosis.

| ORIGA Dataset (Glaucoma) |  |  |  |  |  |  |  |
| --- | --- | --- | --- | --- | --- | --- | --- |
| Method | Nor:Abn | Train:Test | Class | Sen | Spec | Acc | AUC |
| EAMNet [60] | 482:168 | 2 fold CV | 2 | - | - | - | 88.0 |
| SVM-SMOTE [61] | 482:168 | 10-fold CV | 2 | 87.6 | 77.9 | 82.8 | <u>88.9</u> |
| 18-CNN [62] | 482:168 | 70:30 | 2 | 58.1 | 92.4 | 78.3 | - |
| NasNet [63] | 482:168 | 70:30 | 2 | 78.7 | 91.1 | 87.9 | - |
| CNN-SVM [64] | 660:453 | 70:30 | 2 | <u>89.5</u> | **100** | <u>95.6</u> | - |
| ODGNet [65] | 482:168 | pretrained | 2 | **94.8** | 94.9 | **95.8** | **97.9** |
| **Topo-ML** | 482:168 | 80:20 | 2 | 50.0 | 92.7 | 81.5 | 77.3 |
| **Topo-Net** | 482:168 | 85:15 | 2 | 84.6 | <u>95.8</u> | 82.3 | 83.0 |

**TABLE V:** Accuracy results for multiclass DR diagnosis.

| APTOS 2019 Dataset - 5-class (DR) |  |  |  |  |  |  |  |
| --- | --- | --- | --- | --- | --- | --- | --- |
| Method | Nor:Abn | Train:Test | Class | Prec | Rec | Acc | AUC |
| DRISTI [56] | 1805:1857 | 85:15 | 5 | 59.4 | 54.6 | 75.5 | - |
| C-DNN [57] | 1805:1857 | 85:15 | 5 | - | - | <u>80.9</u> | - |
| SCL [59] | 1805:1857 | 85:15 | 5 | <u>73.8</u> | 70.50 | **84.6** | <u>93.8</u> |
| **Topo-ML** | 1805:1857 | 80:20 | 5 | 73.5 | <u>75.6</u> | 75.6 | 88.3 |
| **Topo-Net** | 1805:1857 | 85:15 | 5 | **82.2** | **78.2** | 80.0 | **95.5** |

In the multiclass setting, again our basic model is handling this challenging 5-class classification problem with simple ML classifier. On the other hand, our Topo-Net model again outperforms all existing baselines in multiclass DR diagnosis.

#### Glaucoma Detection Results

We give our results for Glaucoma diagnosis on ORIGA dataset in Table IV. Among these three retinal diseases, we get our worst performance in Glaucoma detection. Our Topo-ML model falls short of the performances of the SOTA deep learning models. Similarly,

**TABLE VI:** Ablation Study. Accuracy results of our Topo-ML model with different subsets of feature vectors for the default setting for each dataset as described in Sec.V-C.

| Feature Vector | Features | APTOS |  | IC-AMD |  | ORIGA |  |
| --- | --- | --- | --- | --- | --- | --- | --- |
|  |  | Acc | AUC | Acc | AUC | Acc | AUC |
| Gray (Betti-1) | 100 | 91.95 | 97.01 | 86.25 | 81.17 | 70.00 | 67.87 |
| Gray (Betti-0,1) | 200 | 91.95 | 97.42 | 86.25 | 82.14 | <u>78.46</u> | 74.39 |
| RGB (Betti-0,1) | 600 | <u>94.00</u> | <u>98.03</u> | 87.50 | 84.63 | 72.31 | 68.69 |
| RGB+Gray (Betti-0,1) | 800 | 93.86 | **98.10** | <u>88.75</u> | **92.53** | 73.08 | <u>75.12</u> |
| Feature Selection | * | **94.54** | 97.94 | **91.25** | <u>90.58</u> | **81.54** | **77.33** |

Topo-Net model does not reach its performance in the other two retinal diseases. We would like to add that the baselines in these datasets are specifically focused on Glaucoma detection, and they train their deep-learning models with serious data augmentation and other methods. Our Topo-Net model uses pre-trained deep-learning models.

### D. Ablation Study

In Table VI, we present the results of our ablation study for the Topo-ML model. We generated 800 topological features for each fundus image by employing different dimensions (k = 0, 1) and considering four different color channels (Gray, RGB). For ML classifiers we utilized XGBoost for the APTOS, ORIGA, and IChallenge datasets. To enhance our Topo-ML model’s performance by mitigating collinearity, we implemented a feature selection algorithm. In the case of APTOS 2019, we selected the top 159 features out of 800, while for IC-AMD, we selected the best 58 features out of 800. For ORIGA dataset, we used the top 84 features out of 800.

In Table VII, we present an ablation study for our Topo-Net model. We integrated 10 different pre-trained models as the backbone for our topological deep learning model. Additionally, we provide comprehensive performance metrics for each of these models in Table X (Glaucoma), Table IX (AMD), and Table VIII (DR). Notably, our topological deep learning model demonstrates substantial performance improvements, especially in the cases of Glaucoma and DR multiclass classification. We wish to emphasize that using a pre-trained model and training a deep learning model specifically for certain tasks are distinct approaches, with the latter offering significantly higher performance but requiring weeks of training.

**TABLE VII:** Comparison of AUC results of CNN results with Topo-Net results for different backbones.

| Models | APTOS-Binary |  | IChallenge-AMD |  | ORIGA |  | APTOS-5 labels |  |
| --- | --- | --- | --- | --- | --- | --- | --- | --- |
|  | CNN | T-CNN | CNN | T-CNN | CNN | T-CNN | CNN | T-CNN |
| Resnet50 | 98.28 | **98.82** | 95.48 | **92.25** | 78.50 | **80.54** | 90.54 | **95.51** |
| DenseNet201 | 97.81 | **98.27** | 90.28 | **84.19** | 75.90 | **76.11** | 90.81 | **92.53** |
| InceptionV3 | 95.87 | **95.10** | 87.30 | **84.45** | 82.88 | **83.03** | 89.09 | **91.13** |
| Inc-ResNetV2 | 52.91 | **95.27** | 82.50 | **76.09** | 73.85 | **75.81** | 75.47 | **89.88** |
| MobileNetV2 | 96.72 | **96.85** | 90.56 | **85.42** | 74.71 | **73.27** | 88.80 | **92.31** |
| Xception | 97.47 | **96.50** | 94.71 | **82.75** | 75.90 | **78.49** | 91.00 | **92.14** |
| VGG19 | 97.80 | **97.67** | 91.84 | **91.30** | 76.41 | **82.08** | 95.65 | **95.48** |
| EfficientNetB0 | 98.62 | **98.69** | 94.30 | **88.61** | 77.95 | **80.66** | 92.34 | **95.25** |
| EfficientNetB2 | 98.28 | **98.94** | **94.68** | **92.30** | 67.74 | **81.62** | 94.25 | **93.46** |
| EfficientNetB3 | 98.68 | **98.51** | 94.47 | **93.55** | 82.46 | **79.66** | 92.85 | **94.62** |

**TABLE VIII:** Accuracy results for Topo-Net on APTOS dataset.

| APTOS 2019 Dataset (DR) - Binary |  |  |  |  |
| --- | --- | --- | --- | --- |
| Method | Acc | Prec | Rec | AUC |
| Resnet50 | 96.73 | 96.73 | 96.73 | 98.28 |
| Topo-Resnet50 | 97.95 | 97.95 | 97.95 | 98.82 |
| DenseNet201 | 96.55 | 96.72 | 96.55 | 97.81 |
| Topo-DenseNet201 | 96.36 | 96.19 | 96.36 | 98.27 |
| InceptionResNetV2 | 52.91 | 52.91 | 52.91 | 52.91 |
| Topo-InceptionResNetV2 | 90.36 | 90.09 | 90.91 | 95.27 |
| MobileNetV2 | 94.73 | 94.73 | 94.73 | 96.72 |
| Topo-MobileNetV2 | 94.73 | 94.72 | 94.55 | 96.85 |
| EfficientNetB3 | 96.91 | 97.09 | 96.91 | 98.68 |
| Topo-EfficientNetB3 | 98.36 | 98.36 | 98.36 | 98.51 |
| Xception | 94.73 | 94.74 | 94.91 | 97.47 |
| Topo-Xception | 93.64 | 93.62 | 93.45 | 96.50 |
| VGG19 | 96.91 | 96.91 | 96.91 | 97.80 |
| Topo-VGG19 | 96.91 | 96.21 | 96.91 | 97.67 |
| InceptionV3 | 91.64 | 91.64 | 91.64 | 95.87 |
| Topo-InceptionV3 | 92.55 | 91.50 | 92.00 | 95.10 |
| EfficientNetB0 | 97.64 | 97.64 | 97.64 | 98.62 |
| Topo-EfficientNetB0 | 97.82 | 97.82 | 97.82 | 98.69 |
| EfficientNetB2 | 96.91 | 96.90 | 96.73 | 98.28 |
| Topo-EfficientNetB2 | 98.73 | 98.73 | 98.73 | 98.94 |

**TABLE IX:** Accuracy results for our Topo-Net models on IChallenge dataset with different backbones.

| IChallenge Dataset (AMD) |  |  |  |  |
| --- | --- | --- | --- | --- |
| Method | Acc | Prec | Rec | AUC |
| Resnet50 | 87.34 | 87.34 | 87.34 | 86.54 |
| TDA + Resnet50 | 86.08 | 86.08 | 86.08 | 85.99 |
| DenseNet201 | 78.48 | 79.49 | 78.48 | 88.05 |
| TDA + DenseNet201 | 81.01 | 81.01 | 81.01 | 80.53 |
| InceptionResNetV2 | 21.52 | 21.52 | 21.52 | 50.00 |
| TDA + InceptionResNetV2 | 77.22 | 77.50 | 78.48 | 77.57 |
| MobileNetV2 | 86.08 | 85.90 | 84.81 | 91.67 |
| TDA + MobileNetV2 | 75.95 | 75.00 | 75.95 | 75.95 |
| EfficientNetB3 | 88.61 | 88.75 | 89.87 | 95.95 |
| TDA + EfficientNetB3 | 92.41 | 92.31 | 91.14 | 93.78 |
| Xception | 84.81 | 84.81 | 84.81 | 88.24 |
| TDA + Xception | 78.48 | 76.92 | 75.95 | 77.14 |
| VGG19 | 86.08 | 86.08 | 86.08 | 92.31 |
| TDA + VGG19 | 88.61 | 88.00 | 83.54 | 86.64 |
| InceptionV3 | 72.15 | 72.15 | 72.15 | 82.73 |
| TDA + InceptionV3 | 79.75 | 79.75 | 79.75 | 80.98 |
| EfficientNetB0 | 86.08 | 85.00 | 86.08 | 90.12 |
| TDA + EfficientNetB0 | 88.61 | 88.61 | 88.61 | 90.91 |
| EfficientNetB2 | 87.34 | 87.34 | 87.34 | 90.24 |
| TDA + EfficientNetB2 | 87.34 | 84.52 | 89.87 | 92.60 |

**TABLE X:** Accuracy results for our Topo-Net models on ORIGA dataset with different backbones.

| ORIGA Dataset (Glaucoma) |  |  |  |  |
| --- | --- | --- | --- | --- |
| Method | Acc | Prec | Rec | AUC |
| Resnet50 | 73.08 | 73.08 | 73.08 | 78.50 |
| Topo-Resnet50 | 79.23 | 74.10 | 79.23 | 80.54 |
| DenseNet201 | 68.46 | 67.72 | 66.15 | 75.90 |
| Topo-DenseNet201 | 76.15 | 76.32 | 66.92 | 76.11 |
| InceptionResNetV2 | 73.85 | 73.85 | 73.85 | 73.85 |
| Topo-InceptionResNetV2 | 75.38 | 67.81 | 76.15 | 75.81 |
| MobileNetV2 | 71.54 | 71.54 | 71.54 | 74.71 |
| Topo-MobileNetV2 | 75.38 | 73.53 | 57.69 | 73.27 |
| EfficientNetB3 | 73.85 | 74.05 | 74.62 | 82.46 |
| Topo-EfficientNetB3 | 80.77 | 75.00 | 76.15 | 79.66 |
| Xception | 65.38 | 65.12 | 64.62 | 75.90 |
| Topo-Xception | 76.15 | 77.48 | 66.15 | 78.49 |
| VGG19 | 69.23 | 68.94 | 70.00 | 76.41 |
| Topo-VGG19 | 78.46 | 78.29 | 77.69 | 82.08 |
| InceptionV3 | 75.38 | 75.59 | 73.85 | 82.88 |
| Topo-InceptionV3 | 82.31 | 76.39 | 84.62 | 83.03 |
| EfficientNetB0 | 70.00 | 70.00 | 70.00 | 77.95 |
| Topo-EfficientNetB0 | 78.46 | 74.29 | 80.00 | 80.66 |
| EfficientNetB2 | 64.62 | 65.89 | 65.38 | 67.74 |
| Topo-EfficientNetB2 | 80.77 | 79.49 | 71.54 | 81.62 |

**TABLE XI:** Accuracy results for our Topo-Net models on APTOS dataset (5-labels) with different backbones.

| APTOS 2019 Dataset (DR) for 5-class |  |  |  |  |
| --- | --- | --- | --- | --- |
| Method | Acc | Prec | Rec | AUC |
| Resnet50 | 76.73 | 77.67 | 75.27 | 90.54 |
| Topo-Resnet50 | 80.00 | 82.22 | 78.18 | 95.51 |
| DenseNet201 | 74.36 | 75.83 | 70.18 | 90.81 |
| Topo-DenseNet201 | 73.64 | 78.98 | 70.36 | 92.53 |
| MobileNetV2 | 71.27 | 73.00 | 69.82 | 88.80 |
| Topo-MobileNetV2 | 75.27 | 78.21 | 69.82 | 92.31 |
| EfficientNetB3 | 76.00 | 76.74 | 74.36 | 92.85 |
| Topo-EfficientNetB3 | 79.64 | 81.94 | 76.73 | 94.62 |
| Xception | 71.09 | 77.63 | 64.36 | 91.00 |
| Topo-Xception | 74.55 | 77.64 | 70.73 | 92.14 |
| VGG19 | 78.55 | 86.73 | 71.27 | 95.65 |
| Topo-VGG19 | 78.73 | 83.79 | 72.36 | 95.48 |
| InceptionV3 | 69.45 | 69.91 | 68.00 | 89.09 |
| Topo-InceptionV3 | 72.55 | 79.04 | 63.09 | 91.13 |
| EfficientNetB0 | 78.55 | 78.83 | 78.55 | 92.34 |
| Topo-EfficientNetB0 | 78.18 | 81.91 | 74.91 | 95.25 |
| EfficientNetB2 | 78.55 | 79.81 | 76.91 | 94.25 |
| Topo-EfficientNetB2 | 80.91 | 81.73 | 80.55 | 93.46 |

### E. Discussion

The results presented in Tables II to V illustrate the performance of our models in three of the most prevalent retinal diseases. Notably, our scalable Topo-ML model offers outstanding computational efficiency, capable of processing thousands of images in mere hours. It accomplishes this without the need for pre-training or data augmentation, features commonly employed by other existing baselines in this field. Given these characteristics, our Topo-ML model stands as a highly successful approach for tasks related to AMD and DR diagnosis, delivering results that are exceptionally competitive when compared to state-of-the-art deep learning models.

On the other hand, our Topo-Net model distinguishes itself by not using any training; it solely utilizes pre-trained models. It is worth noting that customizing and training the model specifically for individual retinal diseases, as done with other deep learning models featured in our accuracy tables, could potentially yield even more impressive results. In this research, our primary objective was to emphasize the value of our topological feature vectors and demonstrate their effective applicability in various machine learning models with ease.

## IV. Interpretation of Topological Features

As mentioned in the introduction, one of the main advantages of our model is the interpretability of topological features. In Figures 6 to 8, we illustrate the topological patterns created by normal and abnormal classes for DR, Glaucoma, and AMD diseases, respectively. In these figures, we give median curves and 40% confidence bands of each class for the corresponding dataset. The details of these non-parametric confidence bands and median curves can be found at [66].

**Fig. 6:**
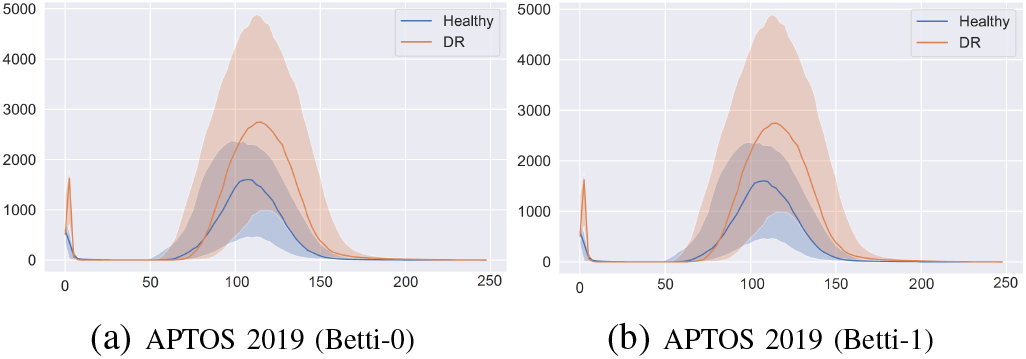
Median curves and 40% confidence bands of our topological feature vectors for DR. *x*-axis represents color values and *β*_0_(*t*) and *β*_1_(*t*) represent the count of components and count of loops in *𝒳*_*t*_, respectively.

To elucidate our model using these figures, the distinct patterns observed in Figures 6 to 8 underscore the discriminative power of our topological feature vectors. From a machine-learning perspective, these visualizations highlight the robustness of our feature vectors. For each image, by using equally distributed 100 thresholds spanning [0, 255] interval, we obtain 100-dimensional Betti-0 and 100-dimensional Betti1 vectors. Consequently, every image is mapped to a point in the latent space ℝ ^100^ through these Betti functions. In this latent space, each image, denoted as **X**, is represented as a point β(**X**) ∈ ℝ^100^, and the median curves can be interpreted as the centroids of clusters corresponding to each class. The separation between these clusters can be thought of as the distance between these points.

**Fig. 7:**
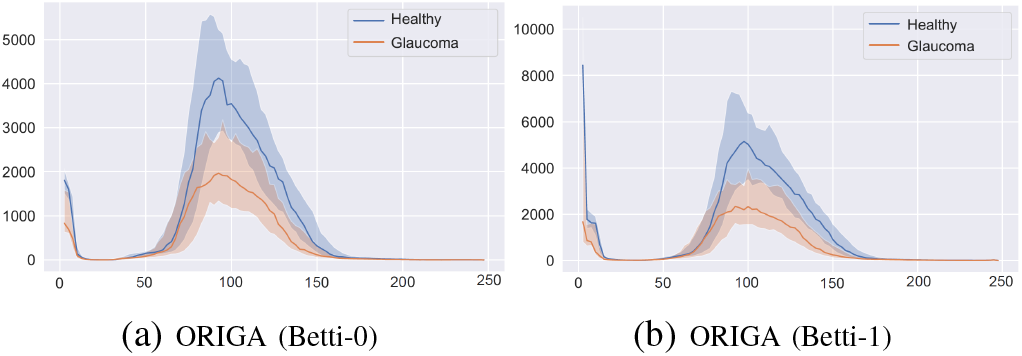
Median curves and 40% confidence bands of our topological feature vectors for Glaucoma. *x*-axis represents color values and *β*_0_(*t*) and *β*_1_(*t*) represent the count of components and count of loops in *𝒳*_*t*_, respectively.

**Fig. 8:**
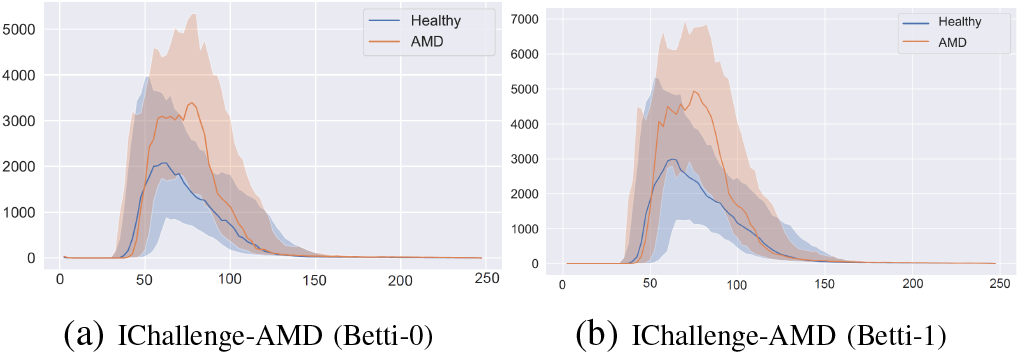
Median curves and 40% confidence bands of our topological feature vectors for AMD. *x*-axis represents color values and *β*_0_(*t*) and *β*_1_(*t*) represent the count of components and count of loops in *𝒱*_*t*_, respectively.

In our visualizations, we provide a highly condensed summary of this latent space. While the curves may appear close to each other, it’s important to note that each entry β_*k*_(t_*i*_) in the vector β_*k*_(**X**) = [ β_*k*_(1) β_*k*_(2) … β_*k*_(100) ] represents the *i*^*th*^ coordinate in ℝ^100^ for an image. Consequently, even a slight separation between the Betti curves results in a substantial distance between the corresponding points in the latent space. This separation between the clusters is readily identified by the machine learning model (e.g., XGBoost) during training, ultimately leading to the development of a highly robust model with exceptional accuracy results.

In order to interpret our features, it’s essential to provide some background on topological feature vectors. While various methods of vectorizing persistence diagrams can yield powerful feature vectors, Betti curves stand out in terms of interpretability. This is one of the key reasons behind our choice to use Betti curves as the vectorization method in our model.

Betti curves keep track of the count of components and loops as we progress through increasing color values from 0 to 255 where the color value 0 represents black, and the color value 255 represents white. In simpler terms, for any color value *t*_0_ ∈ [0, 255], β_0_(*t*_0_) represents the total number of components in the binary image 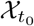, while β_1_(*t*_0_) indicates the total number of loops or holes in the binary image 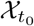 (as illustrated in Figure 3). It’s important to note that the counts on the y-axis of the figures represent the actual counts of components and loops, and they are not normalized values. Keeping this fundamental property in mind, we interpret our topological fingerprints for each disease as follows:

In Figure 6b, we see Betti-1 curves of the two classes (DR vs. normal) in the APTOS 2019 dataset. Our curves indicate that when the grayscale value is between 100 and 150, the count of loops in the DR class is almost double the ones in the normal class. In other words, if *𝒳* is a normal fundus image and *𝒱* is a DR fundus image in the APTOS dataset, the binary image *𝒳*_125_ has around 1000 loops (holes), while the binary image *𝒱*_125_ has around 2200 loops. One way to interpret this is that in a normal class, there are about 1000 light spots in normal classes, while there are about 2200 light spots spread out DR classes as holes in *𝒳*_125_ and *𝒱*_125_ are white regions (color value > 125) in these binary images. Therefore, our results show that in DR fundus images, lighter spots are much more abundant and spread out than in normal fundus images.

Similarly, in Figure 7a, we have Betti-0 curves of the two classes (Glaucoma vs. normal) in the ORIGA dataset. In this case, our curves indicate that when the grayscale value is between 80 and 130, the count of components in the Glaucoma class is almost half of the count of components in the normal class. In other words, if *𝒳* is a normal fundus image and *𝒱* is a Glaucoma fundus image in the ORIGA dataset, the binary image *𝒳*_90_ has around 4000 components, while the binary image *𝒱*_90_ has around 2000 component. One can interpret this as follows: In a normal classes, there are about 4000 dark spots in normal classes, while there are about 2000 dark spots spread out in Glaucoma classes. Less number of components means that Glaucoma images get darker faster than the normal classes. i.e., dark regions get more connected in earlier thresholds.

Finally, in Figure 8b, we see Betti-1 curves of the two classes (AMD vs. normal) in the IChallenge-AMD dataset. Our curves indicate that when the grayscale value is between 50 and 100, the count of loops in the AMD class is almost double the ones in the normal class. In other words, if *𝒳* is a normal fundus image and *𝒱* is an AMD fundus image in the IChallenge-AMD dataset, the binary image *𝒳*_75_ has around 2500 loops (holes), while the binary image *𝒱*_75_ has around 5000 loops. Like DR case, this interpreted density of light spots in the images of the AMD class is much higher than the images in the normal class. Here, grayscale value 50 represents a very dark color and grayscale value 0 represents black.

## VII. Conclusion

In this paper, we introduce a novel approach to retinal image analysis by incorporating topological data analysis techniques. Through the application of persistent homology, we generate highly effective topological feature vectors from fundus images, specifically targeting prevalent retinal diseases like DR (Diabetic Retinopathy), Glaucoma, and AMD (Age-Related Macular Degeneration). These topological features, when combined with simple machine learning classifiers, effectively distinguish between normal and abnormal images for these diseases, yielding competitive accuracy results when compared to existing deep learning models. Furthermore, when we combine these topological features with deep learning models in our Topo-Net framework, we surpass state-of-the-art models in benchmark datasets for DR and AMD.

An additional advantage of our approach is the high interpretability of the topological features, which can provide valuable insights to expert ophthalmologists, aiding in their understanding of the subtleties of these diseases. Moreover, these topological feature vectors, being versatile feature representations, can seamlessly integrate with various machine learning and deep learning models. Given the pressing need for automated clinical decision support systems to assist healthcare professionals, our unique topological feature vectors have the potential to significantly enhance the performance and robustness of future machine learning and deep learning models in this domain.

## Data Availability

All data produced are available online at

https://refuge.grand-challenge.org/iChallenge-AMD/

https://www.kaggle.com/datasets/arnavjain1/glaucoma-datasets

https://www.kaggle.com/datasets/mariaherrerot/aptos2019

## VIII. Appendix

Below, we provide additional performance metrics for our Topo-Net model, utilizing various pre-trained CNN models as backbones.

## Footnotes

1 https://github.com/FaisalAhmed77/Topo-Net

